# Clinical Presentation of Psychotic Experiences in Patients with Common Mental Disorders Attending the UK Primary Care Improving Access to Psychological Therapies (IAPT) Programme

**DOI:** 10.1101/2023.08.26.23294566

**Authors:** Anna Wiedemann, Jan Stochl, Debra Russo, Ushma Patel, Polly-Anna Ashford, Naima Ali, Peter B Jones, Jesus Perez

**Affiliations:** Department of Psychiatry, University of Cambridge, UK; Cambridgeshire and Peterborough NHS Foundation Trust, UK; National Institute for Health Research, Applied Research Collaboration, East of England, UK; Department of Kinanthropology and Humanities, Charles University, Czechia; Norwich Medical School, University of East Anglia, UK; Institute of Biomedical Research, Department of Medicine, University of Salamanca, Spain

## Abstract

**Background:** Improving Access to Psychological Therapies (IAPT) services address anxiety and depression in primary care, but psychotic disorders are typically excluded. Our previous research found that 1 in 4 patients report distressing psychotic experiences (PE) alongside common mental disorders, yet little is known about their clinical presentation and their impact on recovery.

**Methods:** We used the Community Assessment of Psychic Experiences - Positive Scale (CAPE-P15) to determine the clinical presentation of PE within IAPT settings. We identified different classes (sub- groups) based on the reported frequencies of PE within this population using latent class analysis.

**Results:** A total of 2,042 IAPT patients completed the CAPE-P15. The mean age was 39.8 (± 15.3) years. We identified five distinct classes of symptom profiles, findings that PE were common, especially self-referential and persecutory ideas. Prevalence and intensity increased across classes, extending to bizarre experiences and perceptual abnormalities (hallucinations) in the fifth and least common class. Perceptual abnormalities were a strong indicator of symptom severity, and patients reporting such experiences were the least likely to achieve recovery by the end of treatment.

**Limitations:** We collected data from IAPT services that included self-report questionnaires, which may have affected the validity of the reported experiences. We did not collect information on negative PE.

**Conclusions:** Patients seeking treatment for anxiety and depression in primary care commonly experience a wide range of positive PE. Self-referential and persecutory ideation emerged as prevalent experiences; perceptual abnormalities were infrequent. Providing information about prevalence and tailoring therapy may help reduce patient distress.

## 1. Introduction

Psychotic experiences (PE) such as paranoid beliefs are relatively common in the general population (Staines et al., 2022). It is estimated that around 1 in 13 people will have had some form of PE by the time they turn 75 years old; the likelihood, however, declines from early to late adulthood (McGrath et al., 2016). Whilst such experiences are usually transient, for about 20% of people they will recur, yet only about 7% of those with PE will go on and develop a psychotic disorder (Linscott and Van Os, 2013). Nonetheless, people with PE – compared with those without – are more than twice as likely to seek treatment from mental health care services (Bhavsar et al., 2018; DeVylder et al., 2014).

Indeed, the co-occurrence of PE with other non-psychotic mental disorders is well documented. Stochl and colleagues, for instance, demonstrated that psychotic phenomena frequently co-occur with anxiety and depression with PE acting as a marker of severity (Stochl et al., 2015). Further research has also shown that individuals with PE often show increased comorbidity, suicidality, and poorer treatment outcome (Healy et al., 2019; Wigman et al., 2012; Yates et al., 2019).

Most research examining the impact of PE focuses on identifying people who are at increased risk for developing psychotic disorders, usually examined within specialised secondary care settings (e.g., Fusar-poli et al., 2012; Hui et al., 2013), or on studying psychosis as a symptom of vulnerability to mental disorders in the general population (e.g., Varghese et al., 2011). Conversely, there is far less research examining the prevalence and impact of PE on help-seeking individuals within primary care settings. These services, however, are often the first point of contact for those experiencing mental health problems. In the UK, about 90% of adults with mental health issues are supported at primary care level through the National Health Service (NHS; NHS Mental Health Taskforce, 2016). Access to psychological therapies significantly increased since the introduction of a national programme in 2008 known as Improving Access to Psychological Therapies (IAPT; https://www.england.nhs.uk/mental-health/adults/nhs-talking-therapies/). The programme was designed to make evidence-based psychological therapies for anxiety and depressive disorders more widely available (Clark, 2018). Psychotic disorders are generally an exclusion criterion and psychotic experiences are not measured. In reality, however, these services are increasingly serving a population with complex and co-morbid conditions (Buckman et al., 2018; Goddard et al., 2015; Hepgul et al., 2016).

We previously hypothesised that PE would be prevalent in the higher tier of IAPT services which treat moderate-to-severe anxiety and depression. We found that at least 1 in 4 people who receive treatment for common mental disorders from these services report PE (Knight et al., 2020; Perez et al., 2018). We measured this using the Community Assessment of Psychic Experiences – Positive Scale (CAPE-P15; Capra et al., 2017, 2013). As predicted, we demonstrated that these patients present with higher initial severity across both anxiety and depression measures, and were less likely to recover by the end of treatment (Knight et al., 2020). Currently, we do not fully understand the clinical presentation of PE within these settings; for instance, we do not know if the full gamut of psychotic psychopathology familiar in psychotic disorders is seen in people presenting with anxiety and depression, whether some experiences are more commonly reported than others, or whether certain phenomena are absent. The knowledge gained could be used to help tailor treatment plans as well as being of theoretical importance for understanding the relationship between anxiety, depression, and PE.

### 1.1 Study Aim

This study assessed the clinical presentation and symptomatic profile of PE within IAPT services and its relationship to anxiety and depression. The study expanded on our previous examination of prevalence and recovery from common mental disorders in the presence of PE (Knight et al., 2020) and is part of a of a wider, innovative UK National Institute for Health Research (NIHR) programme grant for applied research which aims to assess the reconfiguration of existing treatment protocols to assess whether prospects for recovery in primary care patients with PE could be improved (Ashford et al., 2022).

## 2. Methods

### 2.1. Setting

We obtained patient data from IAPT services within three NHS trusts including Cambridgeshire and Peterborough NHS Foundation Trust (CPFT), Norfolk and Suffolk NHS Foundation Trust (NSFT), and Sussex Partnership NHS Foundation Trust (SPFT). Services within these geographical areas cover a total population of more than 4 million people living in diverse socioeconomic conditions ranging from urban, suburban, and highly dispersed rural communities (Ministry of Housing Communities & Local Government, 2019). The 321 English local authority districts involved vary widely in terms of deprivation levels, from very deprived (Hastings, Sussex) to least deprived (South Cambridgeshire, Cambridgeshire). Further information about IAPT services is provided in the supplementary materials.

### 2.2 Measures

Where therapists of participating trusts were interested in measuring PE for clinical and service evaluation purposes, they collected the Community Assessment of Psychic Experiences – Positive Scale (CAPE-P15; Capra et al., 2017, 2013). The CAPE-P15 is a 15-item self-report measure of positive PE derived from the original 42-item CAPE measure (Stefanis et al., 2002). The 15 items of the CAPE-P15 are grouped into three dimensions including persecutory ideation, bizarre experiences, and perceptual abnormalities (see Table 1). Its feasibility and acceptability to identify individuals with such experience within primary care settings has already been confirmed (Perez et al., 2018). The questionnaire measures both frequency and associated distress on two separate 4-point Likert scales, providing a mean per-item score for both scales with higher scores indicating a higher frequency of PE and an increased level of distress associated with such experiences. When calibrated against the comprehensive assessment of at-risk mental states (CAARMS; Yung et al., 2005), a semi-structured assessment tool to identify individuals at-risk mental states (ARMS) for psychosis, a score of 1.47 or higher on the CAPE-P15 indicates clinically significant PE akin to being positive on the CAARMS (Bukenaite et al., 2017). We will refer to these patients as being CAPE-P15 positive for the purpose of this article.

**Table 1.** Endorsement of the CAPE-P15 items (*n* = 2,042).

| Items | Never | Sometimes | Often | Nearly always | Missing |
| --- | --- | --- | --- | --- | --- |
| *In the past 3 months, have you ...* | % | % | % | % | % |
| **Persecutory ideation** |  |  |  |  |  |
| 1. (...) felt as if people seem to drop hints about you or say things with a double meaning? | 30.07 | 41.43 | 21.30 | 7.00 | 0.20 |
| 2. (...) felt as if some people are not what they seem to be? | 26.30 | 38.79 | 25.02 | 9.55 | 0.34 |
| 3. (...) felt that you are being persecuted in anyway? | 53.23 | 30.02 | 11.90 | 4.70 | 0.15 |
| 4. (...) felt as if there is a conspiracy against you? | 69.20 | 20.91 | 5.83 | 3.97 | 0.10 |
| 5. (...) felt that people look at you oddly because of your appearance? | 71.60 | 17.48 | 6.56 | 4.16 | 0.20 |
| **Bizarre experiences** |  |  |  |  |  |
| 6. (...) felt as if electrical devices such as computers can influence the way you think? | 39.91 | 33.50 | 13.91 | 12.44 | 0.24 |
| 7. (...) felt as if the thoughts in your head are being taken away from you? | 77.28 | 14.84 | 5.48 | 2.15 | 0.24 |
| 8. (...) felt as if the thoughts in your head are not your own? | 69.49 | 20.27 | 7.69 | 2.50 | 0.05 |
| 9. (...) thoughts ever been so vivid that you were worried other people would hear them? | 80.22 | 13.66 | 4.36 | 1.52 | 0.24 |
| 10. (...) heard your thoughts being echoed back at you? | 76.59 | 16.85 | 4.55 | 1.71 | 0.29 |
| 11. (...) felt as if you are under the control of some force or power other than yourself? | 81.34 | 12.78 | 3.77 | 1.96 | 0.15 |
| 12. (...) felt as if a double has taken the place of a family member, friend or acquaintance? | 84.72 | 10.09 | 3.57 | 1.37 | 0.24 |
| **Perceptual abnormalities** |  |  |  |  |  |
| 13. (...) heard voices when you are alone? | 92.51 | 4.41 | 1.76 | 0.78 | 0.54 |
| 14. (...) heard voices talking to each other when you are alone? | 94.17 | 3.77 | 1.13 | 0.69 | 0.24 |
| 15. (...) seen objects, people or animals that other people can't see? | 88.98 | 8.28 | 2.15 | 0.34 | 0.24 |

Routine measures collected in IAPT services included the generalised anxiety disorder questionnaire (GAD-7; Spitzer et al., 2006) and the patient health questionnaire (PHQ-9; Kroenke et al., 2001). In general, services use several improvement metrics to assess recovery from anxiety and depression, most commonly, however, they use the recovery index established by Gyani and colleagues (Gyani et al., 2013). This index states that a patient is considered recovered if they score above the clinical cut-off on the GAD-7 (8 or more points) and/or the PHQ-9 (10 or more points) at the beginning of treatment (within these services known as ‘caseness’), show reliable improvement during treatment, and score below these clinical cut-offs on both the GAD-7 and PHQ-9 after treatment finished. The percentage of patients who recover after accessing IAPT services varies significantly across England, with 50% recovering nationally in 2021/22 (NHS Digital, 2022). Reasons for the variation have been attributed to organisational factors, such as the number of sessions received, staff experience, fidelity to a therapeutic model, and variation in the clinical complexity of patients seen across services (Delgadillo et al., 2014; Gyani et al., 2013).

### 2.3 Sample

We obtained data from the entire caseload of each of the three trusts from February to December 2018, however, only a limited number of patients were approached to complete the CAPE-P15 as part of this service evaluation. Patients of participating trusts could be approached by their therapist to complete the CAPE-P15 once during their course of treatment. This may have been at any time deemed appropriate by the therapist. The CAPE-P15 was given to patients with a brief explanation of the study and instructions on its completion. All patients were told that completing the questionnaire was voluntary. CPFT and SPFT offered to complete the CAPE-P15 during a treatment session, or as a homework task. NSFT collected the CAPE-P15 alongside other routine clinical data using a digital portal, but also offered completing the CAPE-P15 together with a therapist.

Our study was approved by and registered with the official NHS Quality Improvement Programmes of all participating NHS Foundation Trusts and confirmed as such by the UK Health Research Authority (https://www.hra.nhs.uk/). We followed the UK Anonymisation Standard for Publishing Health and Social Care Data (https://digital.nhs.uk/) guidelines for data analysis.

### 2.4 Statistical Analysis

Prevalence of PE was determined using a cut-off of 1.47 for both sub-scales, frequency and associated distress, of the CAPE-P15 (see section 2.3 on measures). Recovery prevalence was calculated for patients who had been discharged from the service according to the recovery index by Gyani and colleagues (2013). The analysis sample consisted of patients who had at least two treatment sessions, not counting any triage sessions, and attended at least one appointment after completing the CAPE-P15.

We used latent class analysis (LCA) to identify sub-groups (*aka* classes) of qualitatively different PE profiles. LCA is a statistical technique used to test whether the observed data are best described as a model that assumes the existence of latent classes, or whether the observed data are best described as one class or group, i.e., with no latent structure (null hypothesis) . If the data fit significantly better with the former model, then the latter is rejected, and it is concluded that the data support the existence of different latent classes. To detect different latent classes, we used participants’ responses to the frequency scale of the CAPE-P15 (*Note*: A patient would only complete the CAPE-P15 distress scale if the associated frequency item is endorsed, hence, the focus on the former scale for the purpose of this analysis). Unlike other clustering methods, LCA does not assign individuals to classes on a definitive basis, but rather uses a stochastic approach to make these assignments.

We used the software package poLCA (Linzer and Lewis, 2011) implemented in the R statistical computing environment to estimate latent class models. It uses a modified expectation-maximisation (EM) algorithm with a Newton-Raphson step for parameter estimation (Bandeen-Roche et al., 1997). As this algorithm is sensitive to starting values of the estimator, we estimated each model 20 times with different starting values to ensure stable convergence for final model estimates. It is important to note that the estimated latent classes are unordered categories, hence, the numerical order of the latent classes in the model output is determined solely by the start values of the EM algorithm. To ease interpretation, we ordered classes by increasing proportion of patients scoring above 1.47 on the CAPE-P15, for instance, the first class would comprise no (or very few) CAPE-P15 positive patients, but this would subsequently increase across classes with the final class including the biggest proportion of CAPE-P15 positive patients. We used Akaike’s information Criterion (AIC) and Bayesian Information Criterion (BIC) to determine the optimal model fit to the data with respect to the number of classes. We further computed model entropy which provides a diagnostic statistic indicating how accurately the model identifies classes. Whilst there is no agreed upon cut-off value, a value close to 1 is seen as ideal and values above 0.8 as acceptable (Weller et al., 2020).

All analyses were conducted in R [Version 4.2.1; R Core Team, 2022] as well as MPLUS [Version 8.8; Muthén and Muthén, 2022]; the latter to retrieve model entropy.

## 3. Results

We obtained 2,042 CAPE-P15 questionnaires from patients receiving treatment from participating IAPT services between February and December 2018. This reflects 7% of the entire IAPT caseload of this period. We previously reported sociodemographics characteristics as well as overall prevalence of PE within this sample in Knight and colleagues (2020). For context and ease of readability, however, we briefly report this information again below.

### 3.1 Descriptive Analysis

#### 3.1.1 Sociodemographic Characteristics

As only a proportion of the entire caseload completed the CAPE-P15, we determined whether there were any differences between patients who did and did not complete the CAPE-P15 by analysing differences across age, sex, and ethnicity (*cf*. Table 2). As previously reported, average age of patients who completed the CAPE-P15 (*M* = 39.8 years, *SD* = 15.3) did not differ from the average age of patients who did not complete the CAPE-P15 (*M* = 39.2, *SD* = 15.3). Similarly, we found no differences in ethnicity across patients who did and did not complete the CAPE-P15. A higher percentage of women (68.9%) compared with men (31%) completed the CAPE-P15.

**Table 2:** Comparison of age, sex, and ethnicity by CAPE-P15 status.

|  |  | CAPE -P15 Status (%) |  |  |
| --- | --- | --- | --- | --- |
|  |  | Positive | Negative | None |
| Age | 17 | 6.4 | 2.8 | 2.7 |
|  | 18-35 | 47.8 | 38.8 | 46.3 |
|  | 36-64 | 44.1 | 50.8 | 44.4 |
|  | 65+ | 1.7 | 7.6 | 6.6 |
| Sex | Male | 29.2 | 32.5 | 32.2 |
|  | Female | 70.8 | 67.5 | 67.8 |
| Ethnicity | White | 85.2 | 90.2 | 89.4 |
|  | Asian, or Asian British | 1.8 | 0.7 | 1.4 |
|  | Black, Black British, Caribbean, or African | 0.8 | 0.8 | 0.7 |
|  | Mixed, or multiple ethnic groups | 3.6 | 0.9 | 1.8 |
|  | Other ethnic group | 0.8 | 0.3 | 0.5 |
|  | Not known | 8.8 | 6.4 | 6.2 |

*Note:* Table as shown in Knight et al. (2020).

#### 3.1.1 Overall Prevalence of Psychotic Experiences

An average of 29.7% (*n* = 606 of 2042) of IAPT patients were CAPE-P15 positive, i.e., scored 1.47 or above on both the frequency and distress scale of the questionnaire. This prevalence, however, differed between the three sites, ranging from 22.5% in CPFT (*n* = 133 of 590), 26.4% in SPFT (*n* = 100 of 379), and 34.8% in NSFT (*n* = 373 of 1073; see Knight et al., 2020) for further details. Across the whole sample, items 1 and 2 were the most commonly endorsed, with about 70% of patients indicating that they had these experiences at least some of the time or more frequently (*cf*. Table 1).

### 3.3 Latent Class Analysis of CAPE-P15 Frequency Items

Using LCA as described in section 2.4, we examined sub-groups based on the frequency items of the CAPE-P15. Table 3 shows model fit indices for a set number of classes where a lower value for both the AIC and the BIC indicates best fit. Whilst the AIC decreased with increasing number of classes, the BIC reached its lowest value for a 5-class solution before increasing again. Fitting more than seven classes resulted in an unstable estimation as well as very small classes of individuals with extreme symptomatic patterns, i.e., outliers. Discordance between AIC and BIC is common in real-world data and stimulation studies have suggested the BIC should be used in such cases (Nylund et al., 2007). The class separation for the 5-class solution was good with an entropy of 0.82.

**Table 3.**
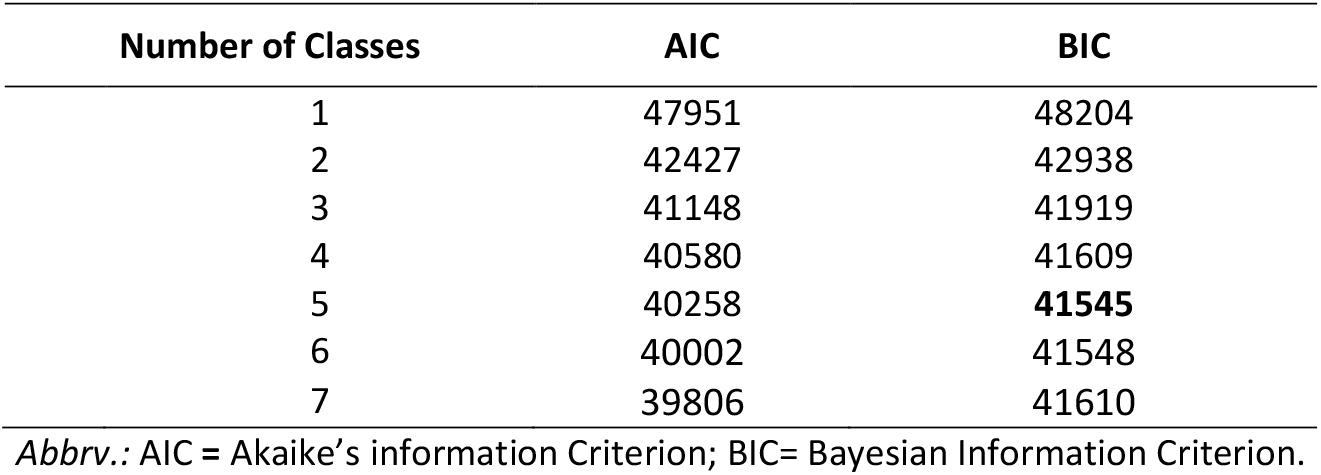
Fit indices for latent class analysis of CAPE-P15 frequency items.

Consequently, we considered the 5-class model as best fit, using these classes for the remainder of the analysis.

#### 3.3.1 CAPE-P15 Status Across Classes

Figure 1 illustrates the proportion of CAPE-P15 positive cases across the five classes. The population share was 19% for Class 1, 36% for Class 2, 20% for Class 3, 21% for Class 4, and 4% for Class 5. Note that each individual has been allocated to the most likely class. Members of Class 1 were all CAPE-P15 negative; almost all of the patients in Class 2 were also CAPE-P15 negative. Class 5, on the contrary, consisted entirely of CAPE-P15 positive patients.

**Figure 1.**
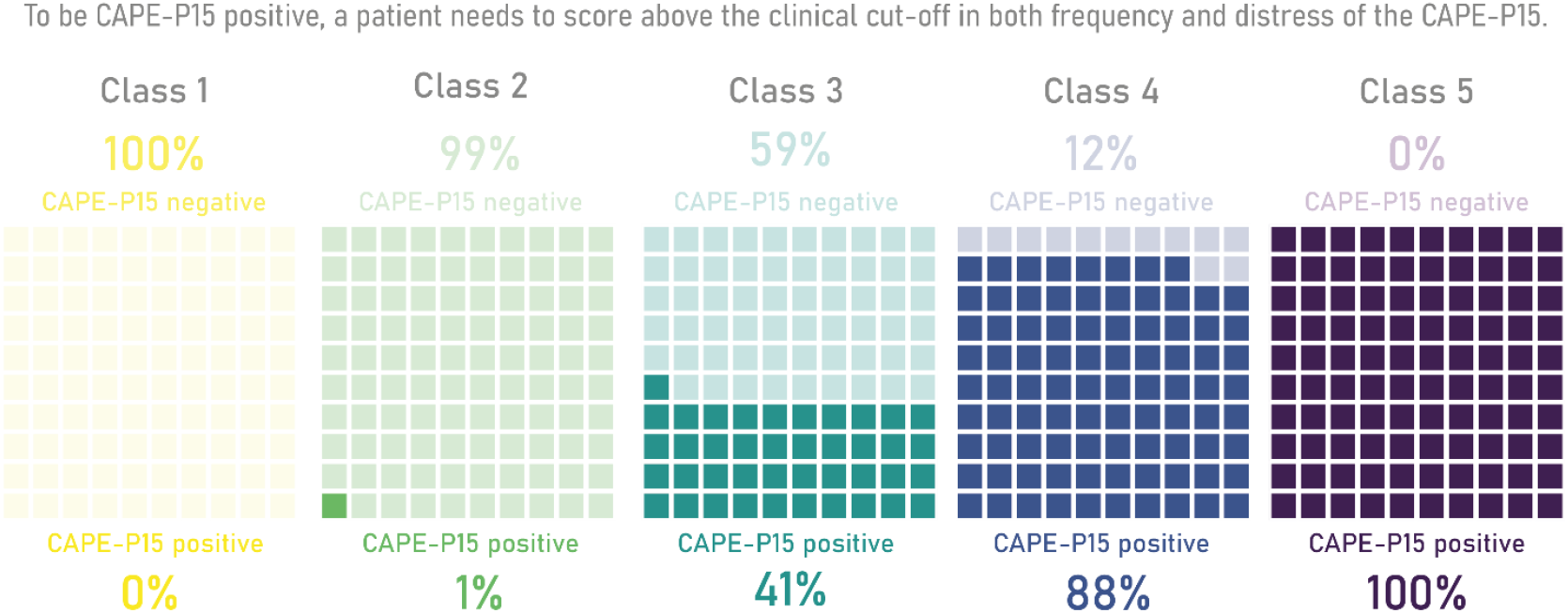
Prevalence of CAPE-P15 negative and CAPE-P15 positive cases across classes. The CAPE-P15 measures psychotic experiences on two 4-point Likert scales, providing a mean per-item score for both frequency and associated distress with higher scores indicating a higher frequency of psychotic experiences and an increased level of distress associated with such experiences. To be considered CAPE-P15 positive, a patient needs to score above 1.47 on both scales frequency and associated distress.

#### 3.3.2 Symptomatic Profile Across Classes

Symptomatic profiles across classes are presented in Figure 2. To ease comparison, we computed weighted mean scores for each CAPE-P15 frequency item across each latent class. A more detailed breakdown of response probabilities as well as further illustrations across classes by individual CAPE-P15 items are provided in the supplementary materials. Whilst patients in Class 1 were most likely to answer all items with “never”, Class 2, the most common, consisted of people who most likely responded sometimes to the first two CAPE-P15 items (“felt people drop hints about me”, and “felt people are not what they seem to be”), but “never” to all other items. Class 3 was relatively similar in profile to Class 2, but PE were slightly more frequent, particularly across the persecutory ideation sub-scale (first five items of the scale) as well as item six which is related to the bizarre experiences sub-scale (“felt electrical devices can influence my thinking”). Class 4 consisted of patients endorsing all CAPE-P15 experiences except perceptual abnormalities. Class 5, whilst the rarest, was the most affected group, with members responding at least sometimes to all frequency questions, including those on the perceptual abnormalities sub-scale (“heard voices when alone”, “heard voices talking to each other when alone”, and “seen objects, people or animals others can’t see”). Overall, items relating to persecutory ideation were the most frequently endorsed, followed by items relating to bizarre experiences; items related to perceptual abnormalities, however, were most likely endorsed by patients in Class 5 (the smallest population with 4%) which exclusively consisted of CAPE-P15 positive patients. Item endorsement across the whole sample can be found in the supplementary materials.

**Figure 2.**
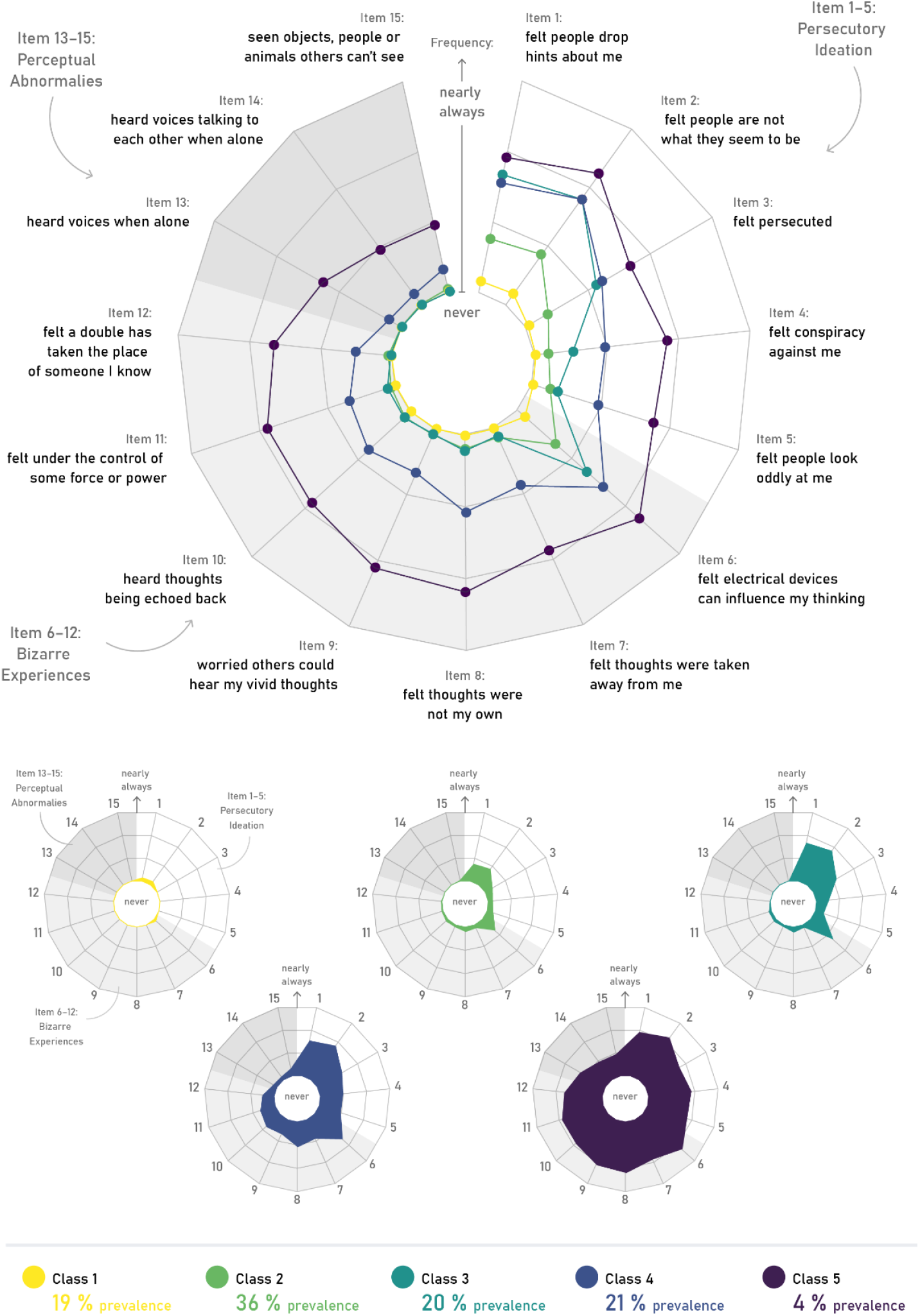
Comparison of psychotic experiences across the three subscales of the CAPE-P15 including persecutory ideation, bizarre experiences, and perceptual abnormalities (top; big radar plot) and psychotic experiences across the CAPE-P15 by class (bottom; smaller radar plots). Please note displayed are expected scores, for response probabilities across all levels of the 4-point Likert scale of the CAPE-P15, see supplementary materials.

**Figure 3.**
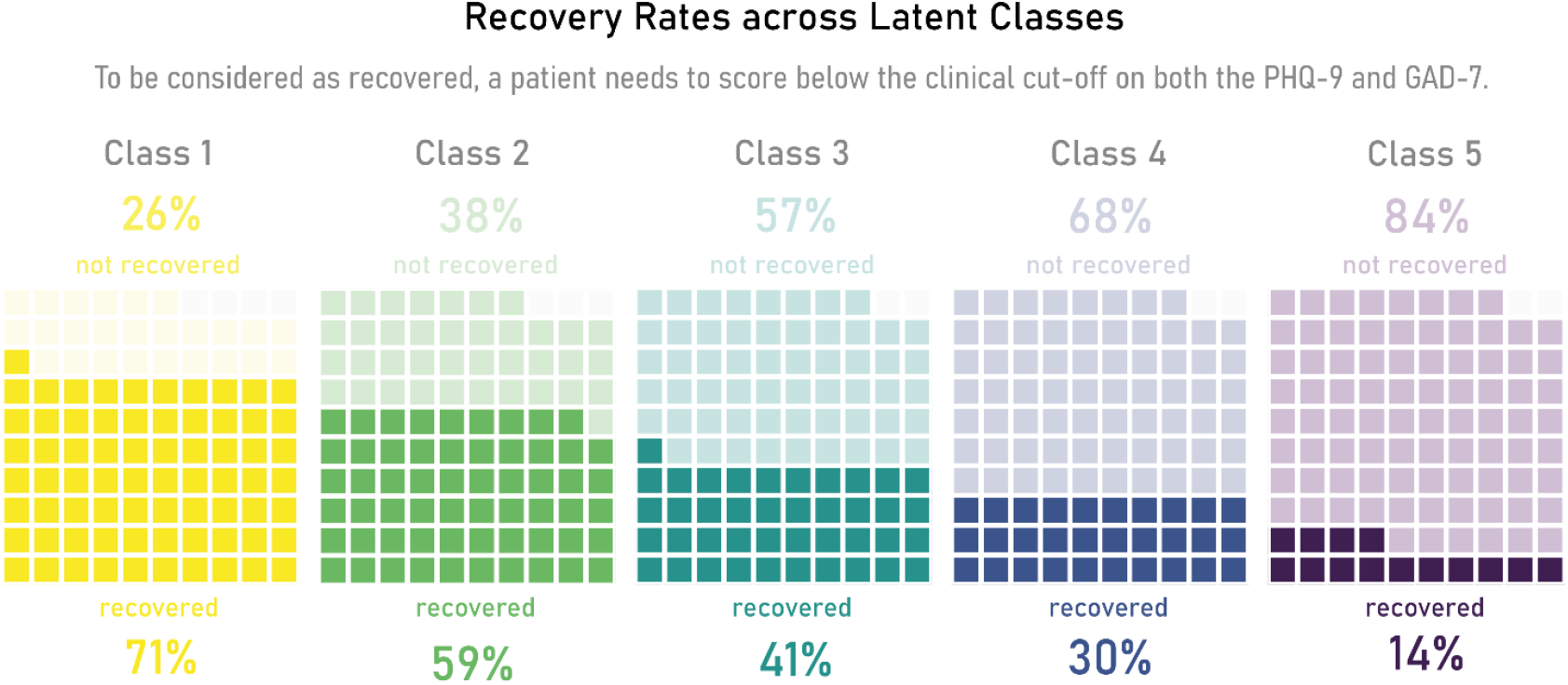
Prevalence of recovery across classes. Under current IAPT guidelines a patient is considered recovered if they score above the clinical cut-off on the PHQ-9 and/or the GAD-7 at the beginning of treatment, show reliable improvement during treatment, and score below the clinical cut-off on both the PHQ-9 and GAD-7 after treatment is finished. Recovery data were missing for 1.6% (*n* = 32) of patients (*n* = 10 for Class 1, *n* = 13 for Class 2, *n* = 4 for each Class 3 and 4, and *n* = 1 for Class 5).

The initial severity for both anxiety and depression increased across classes on average ranging from 11.1 (*SD* = 4.8; Class 1) to 16.9 (*SD* = 3.8; Class 5) for the GAD-7 and from 12.0 (*SD* = 6.0; Class 1) to 20.1 (*SD* = 4.9; Class 5) for the PHQ-9. For reference, clinical cut-offs for the GAD-7 are scores below 8 and for the PHQ-9 scores below 10. Note that initial GAD-7 and PHQ-9 scores were missing for about 12.5% of patients.

#### 3.3.3 Recovery Across Classes

Individuals presenting with a symptomatic profile as seen in Class 1 and 2 were more likely to reach recovery as per current IAPT guidelines compared with individuals in Class 3, 4, and 5. Recovery rates in Class 1 and 2 were above the national average of 50% with 70.7% in Class 1 and 59.4% in Class 2. Individuals in Class 5 were the least likely to recover with only 14.3% reaching recovery by the end of treatment. Recovery data were missing for 1.6% (*n* = 32) of patients (*n* = 10 for Class 1, *n* = 13 for Class 2, *n* = 4 for each Class 3 and 4, and *n* = 1 for Class 5). For further details about recovery rates across the three NHS trusts rather than latent classes, see Knight and colleagues (2020).

## 4. Discussion

The aim of the present study was to assess the clinical presentation of PE in patients with common mental disorders seeking treatment from primary care IAPT services in the UK. We identified five different classes which primarily differed by overall severity of PE. Our results suggest that self-referential and persecutory ideas are particularly common. Around 70% of patients reported feelings such as that people were “dropping hints” about them or that people were “not what they seem to be” at least some of the time. In contrast, PE related to perceptual abnormalities, such as hearing voices when alone, were less common. Nonetheless, rarer experiences always occurred in tandem with self-referential and persecutory ideas.

The high prevalence of self-referential and persecutory ideation may not be surprising given how common such thoughts are in the general population (Freeman, 2007; Staines et al., 2022). Being cautious or attentive to the intentions of others can be beneficial in some situations, however, once these thoughts become excessive or unjustified it can cause significant distress and plausibly this is exacerbated when comorbid with common mental health disorders like anxiety and depression. PE captured in our sample were not restricted to fleeting thoughts that were dismissed almost as they occurred but were significantly distressing for at least 1 in 4 people seeking treatment as reflected by the proportion of patients meeting the clinical cut-off on the CAPE-P15. We further observed that with an increase in the number of PE, there was a higher occurrence of more unusual ideas or experiences, potentially indicative of a hierarchical arrangement of PE (e.g., see Freeman et al., 2005). Our observation aligns with earlier studies that indicate that the most common type of PE is persecutory ideation, followed by unusual experiences that are not based in reality, i.e., bizarre experiences, and abnormal perceptions (e.g., Armando et al., 2010; Capra et al., 2015; Ziermans, 2013). Even though the latter were rare within IAPT settings, they exist and act as a marker of severity; patients endorsing such experiences were the least likely to recover by the end of treatment.

We previously reported that CAPE-P15 positive patients tended to score higher on both the PHQ-9 and GAD-7 at the beginning of treatment. Whilst they followed a similar treatment trajectory as those who were CAPE-P15 negative, they were less likely to achieve recovery by the end of treatment (Knight et al., 2020). In this study, we showed that the PHQ-9 and GAD-7 scores at the beginning of treatment were also associated with the extend of PE in IAPT patients. Specifically, those who reported little to no PE tended to report “only” moderate levels of anxiety or depression. However, as the severity of PE increased, anxiety and depression became more severe. Additionally, recovery rates declined with increasing severity of PE, i.e., only between 14-30% of patients who reported experiences across various CAPE-P15 domains (Class 4 and 5) achieve recovery by the end of treatment, compared with 60-70% of patients that reported minimal or no PE at all (Class 1 and 2). This is consistent with a model whereby PE, at least positive features, are associated with and possibly mediated by affect dysregulation (Wigman et al., 2012).

Altogether, our findings of a five-class solution suggests that the severity of PE might be best represented in (five) levels rather than following a linear pattern which would be warranted if only one class were extracted. As classes primarily differed by overall severity of PE and those with higher PE also reported higher levels of anxiety and depression, our findings indirectly support the existence of a general, transdiagnostic mental distress factor. Transdiagnostic approaches transcend traditional diagnostic boundaries and represent a major change in perspective from conventional diagnostics (e.g., see Caspi and Moffitt, 2018). For instance, Stochl and colleagues demonstrated that psychotic phenomena frequently co-occur with anxiety and depression with PE acting as a marker of severity (Stochl et al., 2015). Our findings suggest PE may indicate the severity of common mental distress rather than the presence of a qualitatively different condition. This would also mean that PE should no longer be seen as a marker of risk for psychotic illnesses such as schizophrenia but rather as part of the network of symptoms that reflect mental ill-health. Nonetheless, we do not know yet know how patients move across the different classes identified as our data can only provide a snapshot. To gain a better understanding, we need longitudinal data that track PE over time.

The high prevalence of PE within primary care settings also offers opportunities for intervention. These may be more effective if we recognise the ubiquity of PE and address the currently underserved needs of this important group of people within primary care settings. Our findings show that individuals with PE are less likely to recover by the end of treatment; only about 14% of patients reporting experiences across the full psychotic spectrum (Class 5) recovered. Even though this sub-group represented only a small proportion of the overall sample, recovery was well below the national average for other patients with distressing, but less severe, PE. Future research should focus on developing interventions that address the specific needs of this group of patients and improve recovery rates. Our current clinical trial TYPPEX is aiming to address at least some of these issues (Ashford et al., 2022). TYPPEX is a multisite, stepped-wedge cluster randomised controlled trial with nested health economic and process evaluations aimed at testing an enhanced training for cognitive behavioural therapists to specifically address PE. The primary objective of the trial is to determine the proportion of patients with common mental disorders and PE who have undergone assessment and treatment and have successfully recovered by the end of therapy.

### 4.1 Limitations

The administration of the CAPE-P15 was not mandatory and only a limited proportion of the entire caseload was offered the assessment. Two of the NHS trusts may or may not have offered the CAPE-P15 as it was agreed that this was at the therapists discretion. A concern could be that therapists may have selected more complex patients which would impact the prevalence estimates of PE. However, the highest prevalence reported was in the trust that distributed the questionnaire automatically.

Furthermore, it is important to acknowledge the limitations of self-report methodologies, which have been well documented in the literature. The discrepancy between the prevalence of PE as assessed through semi-structured interviews and self-report can be as high as 55% (Zammit et al., 2013). For instance, patients may be reluctant to report PE due to social desirability and stigma. However, previous research supports the validity of the CAPE-P15 as a reliable tool for measuring PE (Mark and Toulopoulou, 2016; Núñez et al., 2021). It should also be noted that the CAPE-P15 was not administered at the same point in treatment for all patients. In order to be included in this study, individuals were required to have undergone at least one additional treatment session after the CAPE-P15 was administered. However, further research is needed to gain a more comprehensive understanding of the temporal progression of PE across treatment by collecting longitudinal data. Finally, we focused solely on measuring positive PE, and unfortunately, we were unable to capture the complete spectrum of psychotic experiences that may accompany common mental disorders. Specifically, negative experiences that entail a decline or loss of typical functioning can be more subtle than positive experiences, and therefore demand thorough evaluation.

### 4.2 Conclusions

Patients seeking treatment for common mental disorders often report PE, particularly self-referential and persecutory ideas. We identified five different sub-groups which primarily differed by overall severity of PE. We found that with an increase in the number of PE, there was a higher occurrence of more unusual ideas or experiences. Nonetheless, rarer PE always occurred in tandem with self-referential and persecutory ideas. Although perceptual abnormalities were rare within IAPT settings, they exist and act as a marker of severity; patients endorsing such experiences were the least likely to recover by the end of treatment. Interventions may be more effective if they recognise the ubiquity of PE and address the currently underserved needs of this important group of people within primary care settings.

## Data Availability

All data produced in the present study are available upon reasonable request to the authors.

## Author Statement

Authors PBJ and JP were principal investigators and designed the study. Author AW drafted the first version of the manuscript with the support of DR and NA. Author JS undertook the statistical analysis, had full access to all the data in the study and takes responsibility for the integrity of the data and the accuracy of the data analysis. Author AW reviewed the analysis and created the tables and figures. PA and UP are both involved in the current management of the trial. All authors were involved in the editing of the manuscript and approved its final version.

## Acknowledgements

We are extremely grateful to the IAPT teams who participated in this study and provided access to the required data. We thank the managers and therapists that graciously extended their full cooperation with this evaluation to help us collect the CAPE-P15 data. The study team would also like to acknowledge the contribution of the patients and appreciate their time completing the CAPE-P15. In addition, we thank the Norwich Clinical Trials Unit (NCTU) for its support managing exports of data from the three participating sites. We would also like to thank Friederike von Polenz their support in graphical design.

