## Supplementary Materials for "Clinical Presentation of Psychotic Experiences in Patients with Common Mental Disorders Attending the UK Primary Care Improving Access to Psychological Therapies (IAPT) Programme"

The Improving Access to Psychological Therapies programme, or short, IAPT services, was established in 2008 with the objective to increase the availability of evidence-based psychological treatment for adults with common mental disorders within the National Health Service (NHS) in England. It operates on a flexible referral basis, for instance, allowing individuals to self-refer rather than having to seek a referral from other primary or secondary care services. Since its inception, these services have steadily grown, receiving about 1.8 million referrals and delivering therapy to approximately 1.2 million individuals in 2021/22 (NHS Digital, 2022). The NHS Long Term Plan sets out plans to further expand the programme, aiming to provide access for an additional 380,000 adults per year to reach 1.9 million by 2023/24 (NHS, 2019). Interventions offered are approved by the National Institute for Health and Care Excellence (NICE) and are delivered in line with a stepped-care model, aiming to offer the most effective, but least resource intensive, treatment to each individual. The most frequently offered intervention is cognitive behavioural therapy, but other treatments are also available such as interpersonal psychotherapy, behavioural activation, or couples therapy. The average number of treatment sessions received in 2021/22 was 8, varying in part due to local commissioning guidelines as well as psychopathological severity and clinical judgement.

An integral part of these services is measuring and evaluating service performance, requiring a minimum dataset to track each patient's clinical progress. At the start of treatment as well as at each subsequent session, services are required to record patient-reported outcome measures for depression and anxiety, ensuring that every patient has a clinical endpoint, even if treatment is discontinued unexpectedly. Data are stored using one of two patient management systems, i.e., PCMIS or IAPTUS and submitted to NHS Digital on a monthly basis.

### LCA Supplements

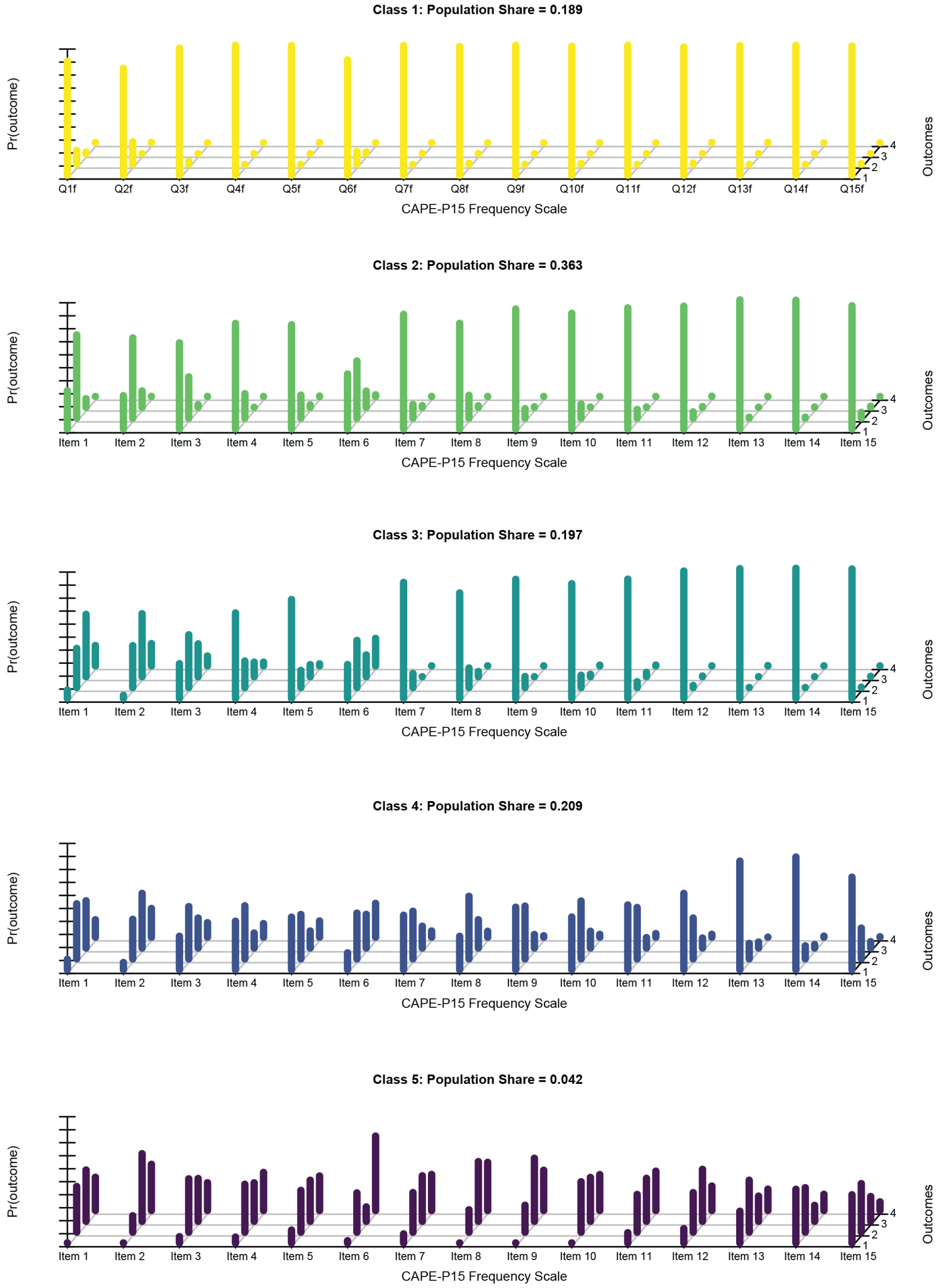


**Figure 1:** Response probabilities and class sizes for a 5-class solution of the CAPE-P15 frequency scale using a finite mixture modelling approach, i.e., latent class analysis. The total population consisted of 2,042 patients who completed the CAPE-P15 whilst receiving treatment at one of the participating talking therapy services. To be included in this study, individuals were required to have undergone at least one additional treatment session after the CAPE-P15 was administered.

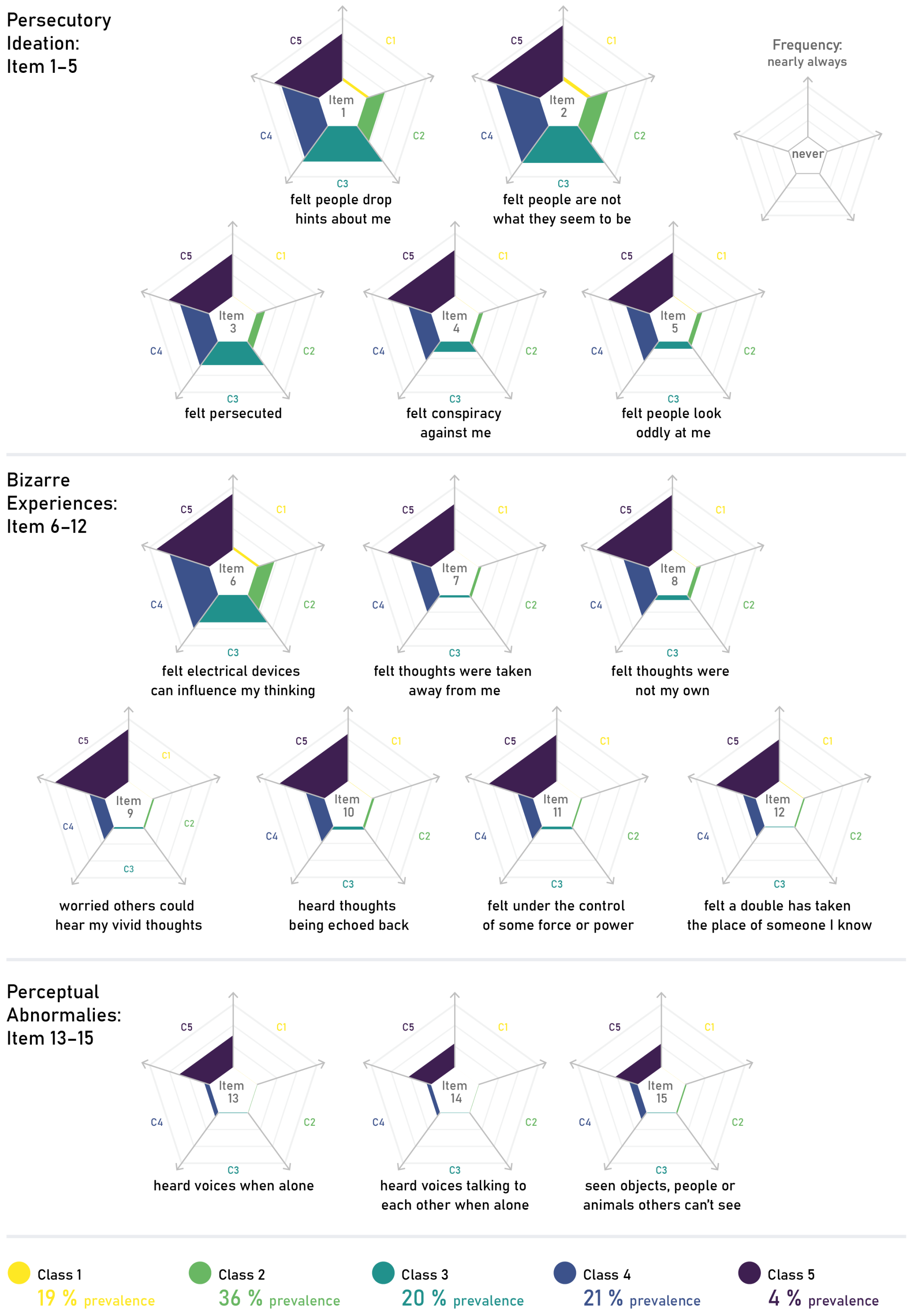


**Figure 2:** Comparison of psychotic experiences across classes and items of the CAPE-P15. Please note displayed are expected scores, for response probabilities across all levels of the 4-point Likert scale of the CAPE-P15, see Figure **??** above.
